# Wide mismatches in the sequences of primers and probes for Monkeypox virus diagnostic assays

**DOI:** 10.1101/2022.08.10.22278644

**Authors:** Fuqing Wu, Jeremiah Oghuan, Anna Gitter, Kristina D. Mena, Eric L. Brown

**Affiliations:** The University of Texas Health Science Center at Houston, School of Public Health Houston, Texas, USA 77030

## Abstract

Rapid and accurate diagnosis of infections is fundamental to containment of disease. Several monkeypox virus (MPV) real-time diagnostic assays have been recommended by the CDC; however, the specificity of the primers and probes in these assays for the ongoing MPV outbreak has not been investigated. We analyzed the primer and probe sequences present in the CDC recommended monkeypox virus (MPV) generic real-time PCR assay by aligning those sequences against 1,730 MPV complete genomes reported in 2022 worldwide. Sequence mismatches were found in 99.08% and 97.46% of genomes for the MPV generic forward and reverse primers, respectively. Mismatch-corrected primers were synthetized and compared to the generic assay for MPV detection. Results showed that the two primer-template mismatches resulted in a ∼11-fold underestimation of initial template DNA in the reaction and 4-fold increase in the 95% LOD. We further evaluated the specificity of seven other real-time PCR assays used for MPV and orthopoxvirus (OPV) detection and identified two assays with the highest matching score (>99.6%) to the global MPV genome database in 2022. Genetic variations in the primer-probe regions across MPV genomes could indicate the temporal and spatial emergence pattern of monkeypox disease. Our results show that the current MPV real-time generic assay may not be optimal to accurately detect MPV, and the mismatch-corrected assay with full complementarity between primers and current MPV genomes could provide a more sensitive and accurate detection of MPV.

## Introduction

Our ability to detect emerging and re-emerging infectious disease agents has never been so important as it is today. While the SARS-CoV-2 is gradually becoming an endemic virus, the world is now seeing the re-emergence of the monkeypox virus (MPV). As of September 26, 2022, there were over 25,000 reported cases in the United States and over 66,000 cases in 106 countries ^1^. The World Health Organization (WHO) has declared a public health emergency of international concern on July 23, 2022 to respond the rapid spread of MPV worldwide.

Monkeypox virus, a species of the *Orthopoxvirus* (OPV) genus, is a zoonotic double-stranded DNA virus with a ∼197 kbp genome ^2,3^. In 1970, monkeypox was first identified in humans in the Democratic Republic of the Congo and in other African nations ^4^. Sporadic monkeypox cases were reported outside of Africa (U.S. and Europe), however, the infections were either related to international travel or exposure to animals imported from Africa ^5,6^. The first MPV case associated with the present outbreak was reported on May 6 in the United Kingdom, followed by cases in Spain, Portugal, and Canada ^5,7^. On May 18, cases were reported in Belgium and Italy; on May 20, cases were reported in Australia, France, Germany, and the Netherlands; and on May 21, cases were reported in Switzerland and Israel ^7^. Such a wide outbreak within two weeks in multiple counties across continents suggests that early non-reported transmission occurred before May 2022 ^8^.

The polymerase-chain reaction (PCR) has been used in the detection and diagnosis of viral infections such as SARS-CoV-2. On June 6, 2022 the Centers for Disease Control and Prevention (CDC) released a real-time PCR test using generic primers and probe for the detection of MPV ^9^ based on previous work conducted in 2010 describing the detection of a West African MPV strain and a Congo Basin MPV strain ^10^. This MPV generic assay has been used to identify human MPV infections and MPV present in wastewater samples ^11,12^. Several other real-time PCR assays have also been reported for MPV or OPV detection in clinical samples ^10,13–17^. However, the specificity and suitability of these generic primers and probes need to be carefully addressed because most of the primers and probes (generic or lineage-specific) were based on sequenced genomes obtained from MPV strains circulating more than a decade ago (2002 to 2009). Given the rapid evolution of viral species, including DNA viruses ^18,19^ it is likely that mutations have emerged in presently circulating MPV strains including the regions targeted by those oligos used for MPV detection. The present report analyzes the oligo sequences used in conjunction with the current real-time MPV assays by aligning primer sequences against 683 reported MPV complete genomes from GISAID as of August 5, 2022. This analysis identified wide variations in the primer and probe sequences compared to the genome of currently circulating MPV stains. Our results indicate that the current MPV real-time generic assay may not be optimal to accurately detect MPV.

## Materials and Methods

### Data source

The MPV genomes were downloaded from GISAID (https://gisaid.org/). As of September 22 (10:50 am), 2022, 1,779 complete MPV genomes were available, including 1,730 genomes (samples) collected and sequenced in 2022. The remaining 49 samples include 7 samples from 1962 to 2005, 11 samples collected in 2006, 12 samples in 2007, 7 samples in 2017, 8 samples in 2018, 2 samples in 2019, and 2 samples in 2021. In the United States, there are 333 complete MPV genomes sampled from May 2022 to August 2022. Metadata includes accession ID, sample collection date, sequence data submission date, and sampling location (continent/country) associated with the sequences. The monkeypox virus reference genome Zaire-96-I-16 was downloaded from NCBI (NC_003310.1). MPV real-time assays (primers and probes) were from published studies including MPV Generic assay ^9^, MPV_O2L ^13^, MPV_F3L ^14^, MPV_C3L ^10^, MPV_G2R_WA ^10^, OPV_B2R ^15^, OPV_I7L ^16^, and OPV_F8L ^17^.

### Sequence alignment

Genome sequences were downloaded as *fasta* files and imported into R (version 4.1.3) as the MPV genome database for analysis. We aligned each of the oligo (including forward primer, reverse primer, and probe) sequences and their reverse complements to the database and computed the percentage of genomes where the oligo sequences were a 100% match. The MPV generic assay and the MPV_G2R_WA assay target two regions of the genome (in inverted terminal repeats region), the other assays have one target. If the oligo is found in more than one target but with different number of matches, the larger value is used as the percentage of 100% match. For example, the forward primer used in the G2R_WA assay is found in sense and antisense strands, but the number of 100% genome matches is 1,779 and 1,778, respectively, therefore, 1,779 was used to calculate the percentage of matching genomes. For non-matches, sequences were extracted and aligned using SnapGene software (https://www.snapgene.com/) to identify the specific nucleotides that differ between the oligos and the genomic data. The major types of sequence variation(s) are listed in Figures 1 and 3A. In total, the original sequence and identified sequence variations accounted for 99.2% ∼ 100% of the total genomes for the MPV real-time assays. These analyses were performed with customized R script.

**Figure 1.**
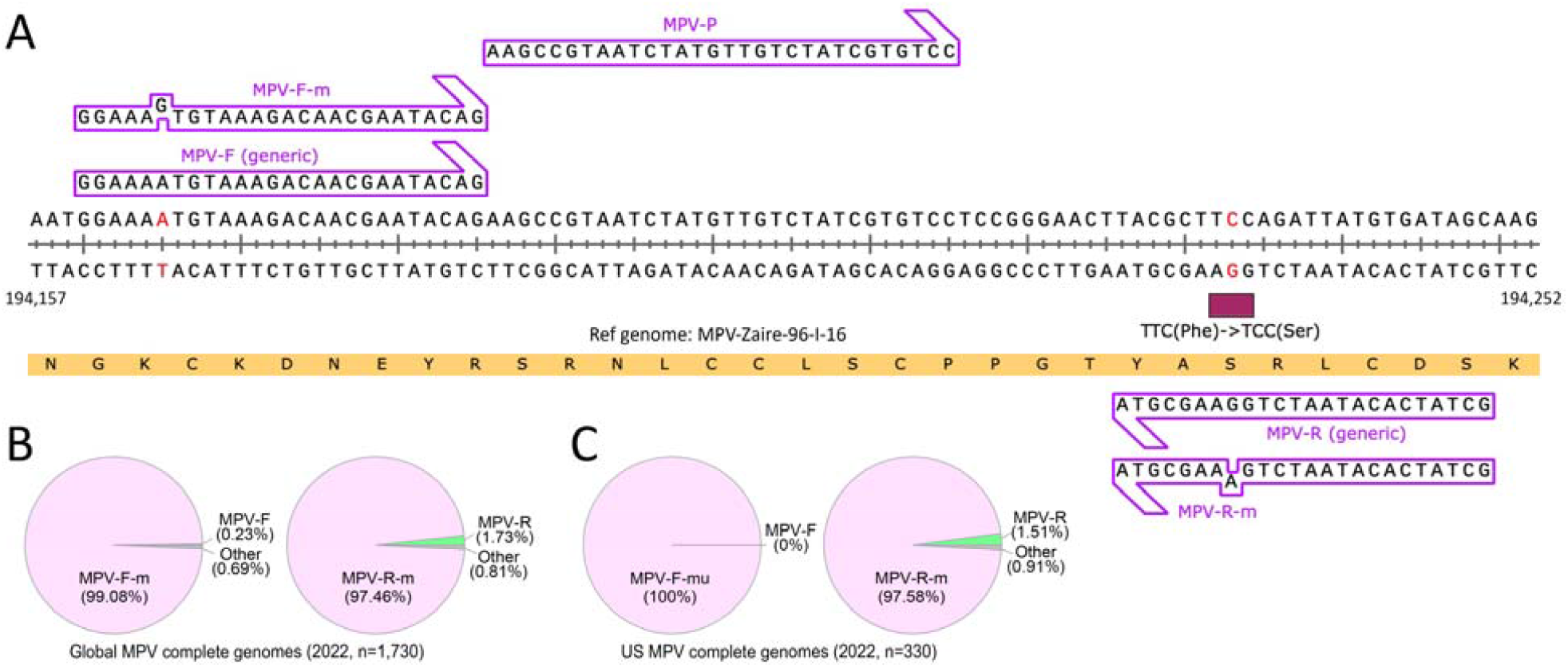
MPV generic forward and reverse primers do not match to the majority of reported genomes. **(A)** Alignment of MPV generic forward primer, reverse primer, and probe sequences (recommended in the CDC test procedure for MPV ^9^) to the MPV reference genome (MPV-Zaire-96-I-16). The corresponding oligo sequences with mismatches found in the genome database for forward (MPV-F-m) and reverse primers (MPV-R-m) were also added on top of the reference genome. Red nucleotides indicate genetic variations identified in the MPV genome database and the nonsynonymous mutation is marketed by a red box. (**B-C**) The abundance of MPV-F (generic) and MPV-F-m, MPV-R (generic) and MPV-R-m in the currently available 1,730 MPV genomes worldwide published in 2022 (B), and in the 330 U.S. genomes (C).

**Figure 2.**
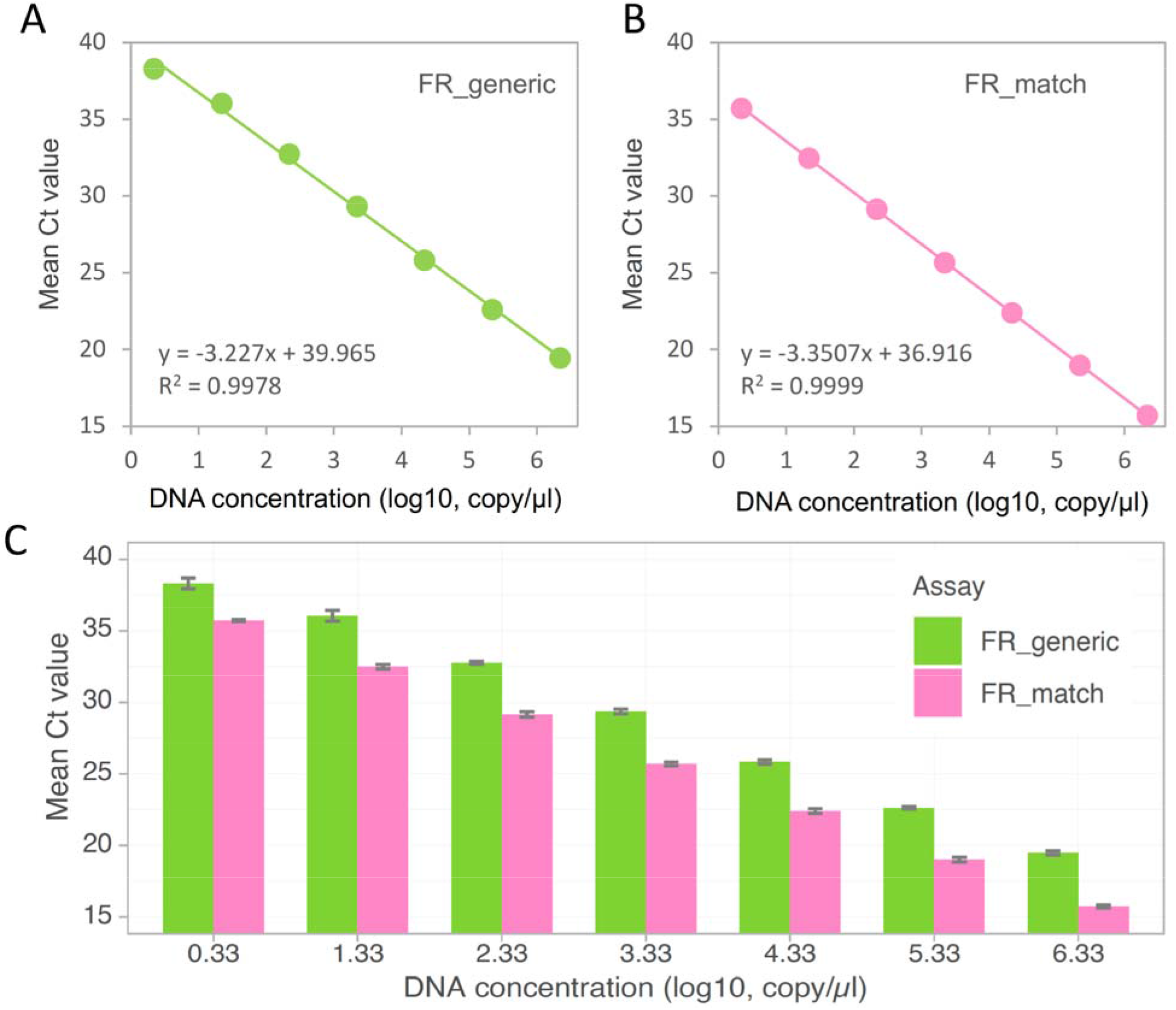
Sensitivity analysis for the MPV generic real time PCR assay and mismatch corrected assay. (**A-B**) Standard curves for MPV FR_generic (A) and FR_match (B) assays. Data shown represent the mean of three replicates with standard deviations. Trendlines with trendline equations and coefficient of determination were added. (**C**) Mismatch-corrected assay (FR_match) has lower Ct values under the same concentrations of initial template DNA.

**Figure 3.**
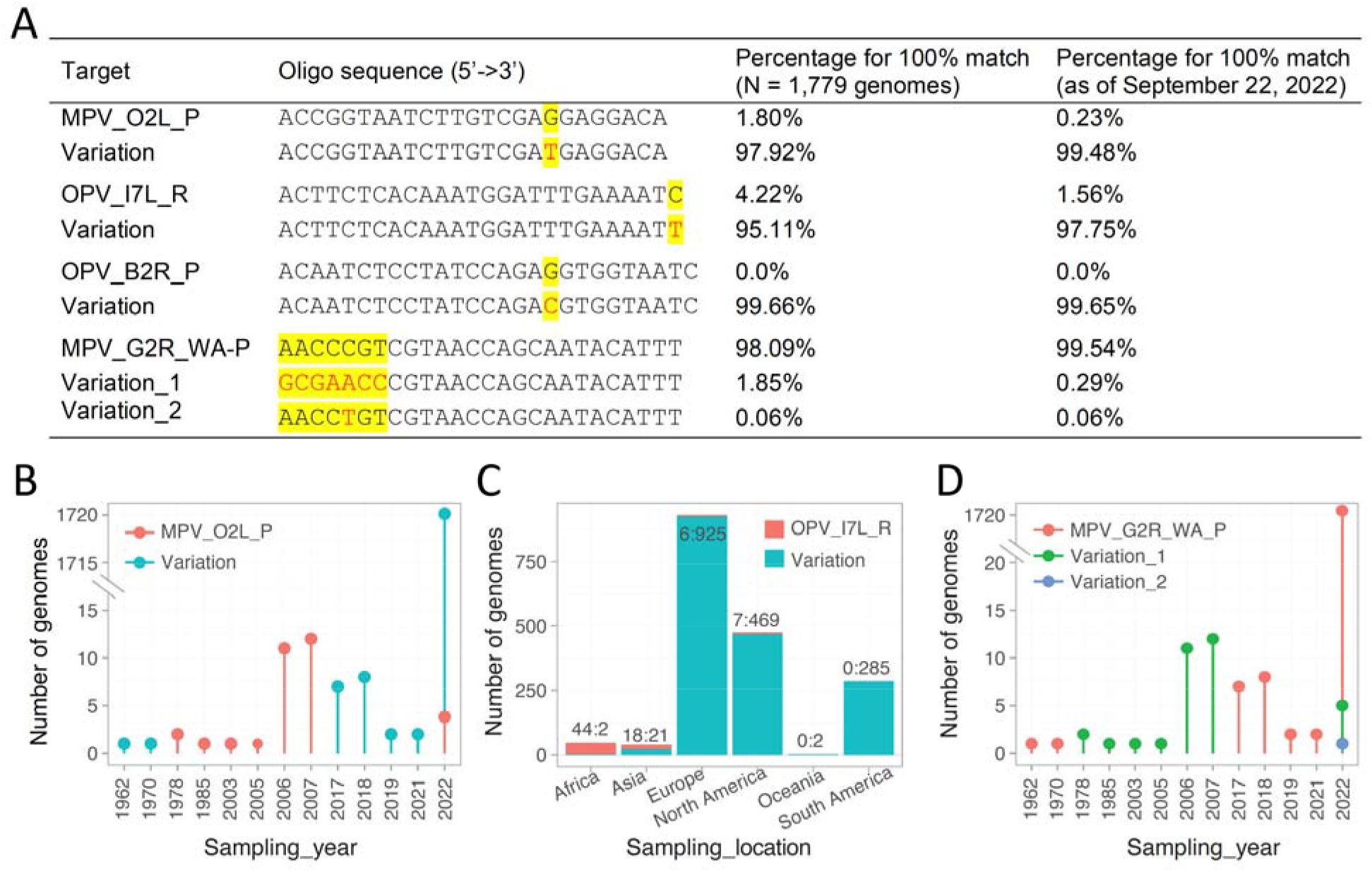
Genetic variations across real-time PCR assays for MPV or OPV detection and epidemiological analysis. **(A)** Genetic variations in the MPV genome database compared to primer and probe sequences used in detection assays. **(B)** Temporal analysis for the genetic variation in MPV_O2L probe sequence. This variation is found in 99.48% of the genomes in 2022. **(C)** Proportion of genomes for the reverse primer used in the OPV_I7L assay with or without variation by continent. **(D)** Incidence of MPV_G2R_WA probe sequence and its two variation types across the genomic database by year.

### Real-time PCR

Quantitative PCR was performed with PCR primers and TaqMan probe. Briefly, the real-time PCR Multiplex Supermix (Bio-Rad, Cat #: 12010220) was mixed with the primers, probe, and nuclease-free H_2_O (VWR, Cat #: 10220-402) and then added to respective wells of a 96-well PCR plate (Bio-Rad, Cat #: HSL9605). For MPV_FR-generic assay, forward primer MPV-F-generic (5’-GGA AAA TGT AAA GAC AAC GAA TAC AG-3’) and reverse primer MPV-R-generic (5’-GCT ATC ACA TAA TCT GGA AGC GTA-3’) were used; for MPV_FR-m assay, mismatch-corrected primers, *i*.*e*., MPV-F-m (5’-GGA AAG TGT AAA GAC AAC GAA TAC AG-3’) and MPV-R-m (5’-GCT ATC ACA TAA TCT GAA AGC GTA-3’), were used (Figure 1A). Probe MPV-P-generic (/56-FAM/AA GCC GTA A/ZEN/T CTA TGT TGT CTA TCG TGT CC/3IABkFQ/) was used for both assays. All primers and probe were synthesized from Integrated DNA Technologies (IDT) and final reaction concentrations were 500 nM and 250 nM, respectively. A 211-bp MPV DNA covering the primer and probe region was synthesized at IDT and used as the PCR template. This synthetic DNA fragment is found in 96.71% (1,673 out of 1,730) of MPV genomes reported in 2022. The standard curves for the MPV generic assay (with mismatches) and for the mismatch-corrected assay (with full complementarity) were generated by serial ten-fold dilutions of the synthesized MPV gene fragment from 2.16*10^0^∼2.16*10^7^ copies/μL. Each concentration has three replicates, and the mean value is used to build the standard curve. For a fine determination of the limit of detection (LOD) for both assays, we used two-fold serial dilutions starting from 21.6 copies/μL and ran 16 replicates at each concentration tested. Another 16 replicates were performed to confirm results for the mismatch-corrected assay at 1.3 copies/μL. Cycle threshold (C_t_) values above 40 were considered negative. The LOD is defined as the lowest concentration of targets at which more than 95% of replicates are positive. The real-time PCR reactions were carried out using a Bio-Rad CFX Opus 96 Detection System at the following conditions: polymerase activation (95 °C for 3 min), PCR (48 cycles, denature at 95 °C for 3 s, and anneal/extend at 60 °C for 30 s).

### Mutation-based epidemiological analysis

For each MPV assay tested, we identified the major variation type(s) in the genome database and then assigned an identity as ‘match’ or ‘variation’ to each genome. Two variation types were found for the MPV_G2R_WA assay and were labeled as ‘Variation_1’ and ‘Variation_2’, respectively. Merging with the metadata associated with the genome, we summarized the number of genomes for ‘match’ and ‘variation’ groups based on the sampling collection time (year), and sampling location (continent). We also sub-grouped the database by only selecting the year ‘2022’ to compute ‘variation’ percentages present in the current MPV outbreak.

## Results

### Generic MPV forward and reverse primers do not match the majority of reported genomes

To investigate whether forward and reverse primers and probe in the MPV generic assay match to the genomes present in the current MPV outbreak, we aligned the three oligo sequences and their reverse complements against the reported 1,730 complete genomes available in 2022. The MPV generic forward sequence (MPV-F) had 100% identity to 4 genomes (0.23%). A single-nucleotide mutation A194165G (synonymous mutation, MPV-F-m in Figure 1A) was found in 99.08% of the published MPV genomes in 2022 (Figure 1B), mostly from May to August 2022. Similar observations were also found in the generic reverse primer (MPV-R), which only matches to 1.73% of the genomes. A nonsynonymous substitution, G194233A in the reverse primer region, was observed in 97.46% of the genomes. In contrast, the probe sequence (MPV-P) is conserved and matches 99.31% of the genomes in the database.

We further compared the MPV generic primers and probe with sequenced samples from the U.S. Of the total of 330 genomes reported from May to August 2022, the A194165G mutation in the forward primer region was observed in all U.S. genomes, and the G194233A mutation observed in the reverse primer was found in 97.58% of the U.S. genomes (Figure 1C) consistent with the results in Figure 1B. These results show that the CDC recommended forward and reverse primers used for MPV generic testing are not 100% identical to the major genome sequences in recent samples with MPV.

### Mismatch-corrected primers have improved quantification and lower limits of detection

Some studies have reported positive detection of MPV in clinical and wastewater samples using the generic real-time assay ^11,12^, suggesting that these primers can amplify the targeted region even with a substitution in each of the primers. We therefore evaluated the real-time PCR performance for the generic primers (FR_generic) and new primers with mismatches corrected (FR_match) using a synthesized 211-bp MPV gene fragment (found in 96.71% of genomes in 2022) covering the primer and probe region. Standard curves from serial 7-log dilutions show that both assays have similar amplification efficiencies with R^2^>0.99 (Figure 2A-B). Under the same concentrations of MPV gene template ranging from 2.16 ∼ 2.16e7 copies/μL, MPV FR_generic has a consistently higher Ct values (3.46±0.39) than FR_match, suggesting that the two mismatches in FR_generic primers cause about 11-fold underestimation of the initial template concentration (Figure 2C). We next evaluated the limit of detection (LOD) by conducting 2X dilutions of template DNA, and each has ∼16 replicates. The 95% LOD for FR_generic assay is 10.8 copies per reaction while the FR_match is 2.7 copies per reaction (Table 1). Together, these results showed that the current MPV generic assay could still detect MPV, but the template concentration quantified from this assay is underestimated by about an order of magnitude. In addition, the mismatch-corrected primers have a lower 95% limit of detection, which is important for samples with low MPV concentrations.

**Table 1.** Limits of detection for MPV generic assay (FR_generic) and mismatch-corrected assay (FR_match).

|  | DNA template volume (μl) | Synthetic MPV gene fragment concentrations (copies / mL) |  |  |  |  |  |  |
| --- | --- | --- | --- | --- | --- | --- | --- | --- |
|  |  | 21,622 | 10,811 | 5,405 | 2,703 | 1,351 | 676 | 0 |
| FR_generic | 2 | 100% (16/16) | 100% (16/16) | 100% (16/16) | 75% (12/16) | 50% (8/16) | Not tested | 12.5% (2/16) |
| FR_match | 2 | 100% (14/14) | 100% (16/16) | 100% (16/16) | 100% (16/16) | 100% (32/32) | 81% (13/16) | 0% (0/16) |

Note: Reactions with Ct < 40 are considered as 'positive'.

### Evaluation of the specificity of other primers and probes for monkeypox virus and orthopoxvirus (OPV) detection

Next, we evaluated seven other developed real-time PCR assays, targeting *O2L, F3L, C3L*, and *G2R* genes for MPV, *B2R, I7L* and *F8L* genes for OPV detection. The selected targets are generally conserved regions within viral genomes to maximize viral detection ^13,15^. We aligned primer and probe sequences and their reverse complements in the seven assays against the total 1,779 MPV genomes and computed the percentage of genomes with a 100% match. As shown in Table 1, the four reported assays designed for MPV detection have varied results. The primers in MPV_O2L have a ∼99% match with the database, but the probe only matches 1.8% of genomes. This ratio goes down to 0.23% when aligned to the genomes in 2022. Primers and probe sequences used in the MPV_F3L assay matches >99.7% of the genomes, suggesting its suitability for detection of the MPV-2022 outbreak. The MPV_C3L assay designed for the detection of the MPV Congo Basin strain poorly matches to the current MPV genomes (0.23%). Primers used in the MPV_G2R_WA assay designed to detect the West African strain showed a 100% match to the database and 99.65% match for the probe, thereby validating previous phylogenetic analysis results that the current MPV lineage belonging to the West Africa clade ^20^.

We further evaluated three real-time assays (OPV_B2R, OPV_I7L, and OPV_F8L) for the detection of viruses belonging to the *Orthopoxvirus* genus, which includes monkeypox virus, camelpox virus, cowpox virus, and vaccinia virus ^21^. OPV_B2R primers highly match the database genomes, but its probe sequence has no match (Table 2). OPV_I7L has good matches for the forward primer and probe, but its reverse primer only matches 1.56% of the genomes. OPV_F8L oligos for the non-variola OPV generic test recommended by CDC has a high match percentage (>99%) with the genomes (Table 2). Thus, the MPV_F3L, MPV_G2R_WA, and OPV_F8L assays matches to >99% of the sequenced MPV genomes.

**Table 2.**
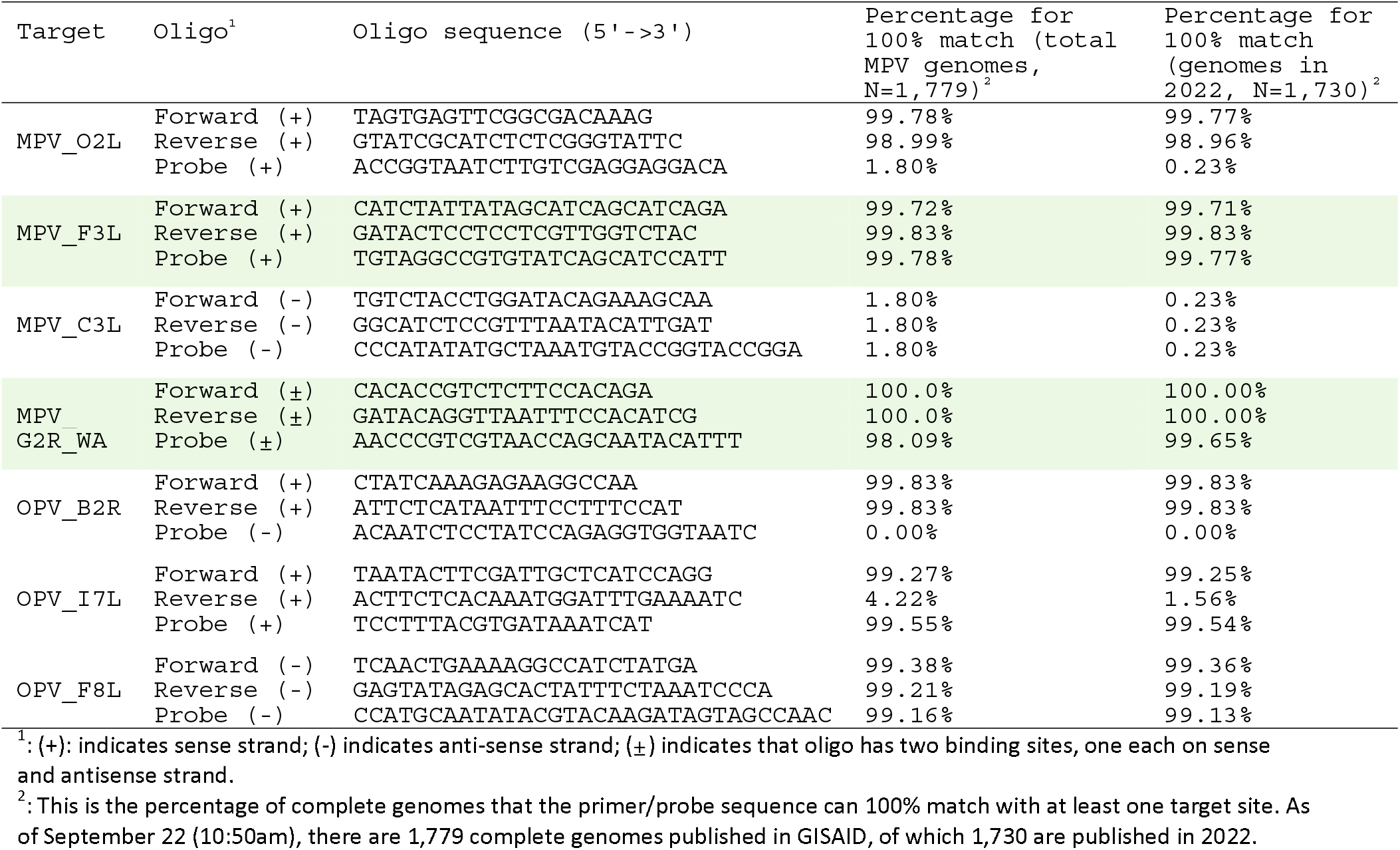
Evaluation of the sequence specificity of primers and probes for monkeypox virus (MPV) or orthopoxvirus (OPV) detection.

### Genetic variations across real-time PCR assays for MPV or OPV detection and epidemiological analysis

Probes in MPV_O2L and OPV_B2R and reverse primer in OPV_I7L match 0 ∼ 1.56% of the genomes, suggesting potential variations in the current MPV genomes. We next analyzed MPV genomic sequences to identify the main variations. About 98% of the MPV genomes have a G->T mutation in the probe used in MPV_O2L assay, and this mutation is present in 99.48% of the MPV-2022 outbreak genomes (Figure 3A). Further analysis based on the sample collection date identified this G->T mutation in two samples in 1962 and 1970, was not detected in samples from 1978 to 2007, and re-emerged from 2017 onward and became dominant in current outbreak (Figure 3B). For the OPV_I7L assay, a C->T mutation was observed at the 3’ end of the reverse primer in 95.11% of all genomes and in 97.75% of genomes sequenced in 2022. It is interesting that this mutation is mostly observed in samples from Europe, South, and North America whereas the original sequence was more common in samples from Africa and Asia (Figure 3C). A G->C substitution in the probe sequence of OPV_B2R was observed in nearly all genomes sequenced in the MPV-2022 outbreak (Figure 3A).

The MPV_G2R_WA assay probe has a 98.09% match to the database sequences (Figure 3A). Two variation types were found for the rest of the 1.91% genomes. A 7-nucleotide variation (variation_1) was found in 28 genomes from 1978 to 2007 (Figure 3A and 3D). Variation_1 reappears in 2022 in one male patient’s sample (age 50) from Luxembourg, and four children samples (age 3 to 10) from Tshopo and Kwilu provinces in the Democratic Republic of the Congo. Although 99.54% of the 2022 MPV genomes are conserved in this probe region, one new C->T synonymous variation (variation_2) (Figure 3A) was identified from a sample obtained on May 18, 2022 from Canada. Those results suggest that the genetic variations in MPV conserved genomic regions covering the real-time PCR primer and probe sequences might be used to indicate the temporal and spatial emergence pattern of monkeypox disease.

## Discussion

Accurate diagnostics are fundamental for successful epidemic control ^22^. PCR-based molecular methods are widely used for pathogen detection in clinical and environmental samples. As a rapid and reliable tool, TaqMan real-time PCR offers high sensitivity and specificity that targets a respective pathogen’s genome by using a designed probe, and forward and reverse primers. Therefore, an exact match between the three oligos and the target genome are required for sensitive and accurate diagnostics. Here, we analyzed the sequence of primers and probe used in the CDC recommended MPV generic real-time PCR assay as well as seven other MPV and OPV diagnostic assays by aligning respective primer/probe sequences against published MPV full genomes. Our results showed two mismatches in more than 99% of genomes for the MPV generic forward and reverse primers (Figure 1). One mutation G194233A is close to the 3’ end of the reverse primer, suggesting it may significantly impact the binding efficiency during amplification.

Theoretically, every mismatch, irrespective of its position within the primer sequence could reduce thermal stability of the primer-template DNA duplex and impact PCR performance ^23,24^. Mismatches have often been reported to undermine real-time PCR based diagnostics. Stadhouders *et al*. examined the impact of primer-template mismatches and found that one G→A mismatch at the 3’end of primer could lead to an increase of >8 Ct values depending on the position ^24^. Klungthong *et al*. found a single mismatch at the 3^rd^ base of the probe significantly reduced sensitivity for detection of influenza A/H1N1/2009 ^25^. In another study, Persson et al reported that that a single G→T mismatch at 3′-end of the reverse primer for detection of hepatitis A virus by RT-PCR assay causes underestimation of the template concentration by a factor of 1.7–1.8 and an ∼2-fold increase in 95% LOD ^26^.

The two identified mismatches, despite neither of them being at the 1^st^ position of 3’ end, may have combined effects on PCR performance. We thus synthesized primers with an identical match to the MPV genome and compared results generated by the assay with mismatches. Our results showed that the two primer-template mismatches resulted in a ∼11-fold underestimation of initial template DNA in the reaction and 4-fold increase in the 95% LOD in our real-time PCR settings. Those results indicated that the mismatch-corrected assay with full complementarity between primers and current MPV genomes could provide a more sensitive detection of MPV, which is important for samples with low viral concentrations such as wastewater ^27–30^. Furthermore, the fully matched primers showed an improved quantification of the initial DNA concentration needed for the quantitative analysis associated with tracking disease progression or assessing estimates of prevalence.

Of the seven other assays evaluated for MPV or OPV detection, MPV_F3L, MPV_G2R_WA, and OPV_F8L had the highest matches (>99%) to the 2022 global genome database. However, specificity, another important parameter for real-time PCR-based diagnostics, needs further investigation for these three assays. For example, we found that primers and probe in the MPV_F3L assay had high a homology to OM460002.1 cowpox virus isolate No-H2 genome (two mismatches in the probe and reverse primer), suggesting it could also amplify cowpox virus. On the other hand, the choice for the three assays may also depend on the sample types. For example, the three assays may work well for clinical samples and provide reliable diagnostics for human MPV. However, extra attention is needed to interpret positive results for wastewater samples, which may contain waste of human, cat, dog, mice, rabbits, and cow origins ^31^.

It is necessary to mention that sequence matching is just one of the parameters determining the detection sensitivity and specificity. Other parameters such as primer dimer formation, GC content, the quality of the template DNA, the PCR master mix, and inhibitors present in the sample may also impact the diagnostic efficiency and mismatch-induced effects ^24,32,33^. An ideal diagnostic assay needs to consider and assess those factors in a comprehensive way. This study used a synthetic MPV gene fragment to test the mismatch effects, but it is recommended to verify the results using a full-length viral genome, which may introduce more complexity and even increase the underestimation ^26^.

In summary, our analysis showed that primers and probe in the recommended MPV generic real-time assay harbor wide deviations from the sequenced MPV genomes, especially the genomes released in current monkeypox outbreak. The mismatch-corrected assay, harboring full complementarity to >97% MPV genomes, shows a higher sensitivity and improved quantification and may be an alternative approach for the MPV detection. Improved diagnostic capabilities are critical as public health practitioners and clinicians respond to the growing MPV outbreak.

## Data Availability

All data produced in the present study are available upon reasonable request to the authors

## Declaration of Competing Interest

The authors declare no competing interest.

## Code Availability

All data and code produced in the present study are available upon reasonable request to the authors.

## Acknowledgement

We are grateful to Dr. Alanna Morrison and the colleagues in the TEPHI for helpful discussion. This work is supported by Faculty Startup funding from the Center of Infectious Diseases at UTHealth, the UT system Rising STARs award, and the Texas Epidemic Public Health Institute (TEPHI).

## Ethical Approval Statement

This article does not contain any studies involving animals or humans performed by any authors.

## Notes

### Competing Interest Statement

The authors have declared no competing interest.

### Summary of Updates

We provided experimental data (Fig 2 and Table 1) to validate bioinformatic analysis.

